# A Pilot Project Leveraging Large Language Models for Automated Screening and Variable Extraction in Observational Studies

**DOI:** 10.64898/2026.06.13.26355589

**Authors:** Manjil M. Pradhan, Rajesh Upadhayaya, Sarah C. Wenyon, Alexandria Viszolay, Melissa Rethlefsen, Vincent Metzger, Gerardo Villarreal, Santiago Alvarez Lesmes, David Andrade, Jason Timm, Scott A. Malec

## Abstract

**Background:** Systematic reviews of observational studies are central to causal inference in chronic disease epidemiology but are increasingly limited by the scale of the literature and heterogeneity in confounder control. There is a need for transparent, open methods that reduce screening burden and make reported exposures, outcomes, and covariates comparable across studies.

**Objective:** To develop and evaluate modular LLM-based pipelines, LitScreen and VarEx, that automate study screening and variable extraction for observational systematic reviews across multiple use cases, including hypertension as a primary exposure with Alzheimer’s disease and related dementias (ADRD) as outcomes, and posttraumatic stress disorder (PTSD) as the exposure with self-harm, self-injury, and suicidality as outcomes.

**Methods and Materials:** We built an end-to-end workflow in which reproducible MEDLINE via Ovid queries yield RIS corpora that are processed by LitScreen, a three-phase screening pipeline combining abstract-level evidence extraction, criterion-wise inclusion adjudication with high-recall gates, and full-text retrieval-augmented verification. Screened-in articles enter VarEx, a retrieval-augmented extraction pipeline that identifies role-specific passages and performs evidence-grounded extraction and semantic classification of exposures, outcomes, and covariates into predefined categories aligned with Metaconfoundr. Performance was evaluated on six labeled SYNERGY datasets and expert-annotated hypertension-to-ADRD and education-to-dementia corpora using precision, recall, F_1_, a strict score requiring correct variable identity and category, and time-per-reference estimates.

**Results:** In the primary hypertension-to-ADRD reference set, VarEx achieved covariate-level precision of 0.80, recall of 0.79, and F_1_ of 0.76, with classification accuracy of 0.97 and similar performance for education-to-dementia and SYNERGY validation datasets. LitScreen preserved high recall while excluding most ineligible records and reduced total screening and extraction time by roughly 80–90 percent relative to manual review baselines by routing only uncertain or borderline citations to full-text verification.

**Conclusion:** A retrieval-augmented LLM framework can automate major components of screening and variable extraction for observational systematic reviews, generating reusable structured covariate inventories that integrate with causal confounder assessment tools and substantially improve the efficiency and reproducibility of evidence synthesis, while remaining an assistant to, rather than a replacement for, human reviewers.

## 1 INTRODUCTION

Observational studies are essential for understanding the multifactorial nature of many chronic conditions, as randomized controlled trials are often infeasible given their chronic nature. Systematic reviews of observational epidemiologic studies provide, when conducted rigorously, the highest level of evidence within non-experimental research because they use explicit, reproducible methods to identify, appraise, and synthesize the totality of available data, thereby supporting more robust causal inference and evidence-based decision making[1].

However, the exponential growth of the biomedical literature has made conducting systematic reviews increasingly resource intensive and time consuming; manual screening of thousands of titles and abstracts often requires many months of effort and substantial personnel time, even for a single review.[2, 3] Empirical studies estimate that contemporary systematic reviews commonly require hundreds to more than one thousand person-hours and a median of more than 60 weeks from protocol registration to publication, with study selection and data extraction representing the most labor-intensive steps.[2, 3, 4, 5] In the context of accelerating publication rates and finite methodological expertise, these demands threaten the timeliness, scalability, and sustainability of evidence synthesis.[3, 5]

These reviews are further challenged by the need to rigorously control for confounding in the underlying observational studies they summarize. Inadequate adjustment, whether through omission of true confounders or inclusion of inappropriate variables such as colliders, mediators, or instrumental variables, can introduce or exacerbate bias, distort effect estimates, and undermine causal interpretation.[6, 7] Principles based on causal diagrams, specifically directed acyclic graphs, and structured confounder selection criteria such as the disjunctive cause criterion have been proposed to guide variable selection and reduce subjective or ad hoc modeling decisions.[6] Nonetheless, empirical assessments of published epidemiologic studies consistently document heterogeneity and inconsistency in the identification and handling of confounders, even for the same exposure–outcome relationships.[6, 8, 9] Figure 1 illustrates the role of a confounding variable in relation to an exposure and outcome of interest, and contrasts this with other common third variable structures such as precision variables, colliders, instrumental variables, and mediators that have distinct implications for adjustment and causal effect estimation.

**Figure 1:**
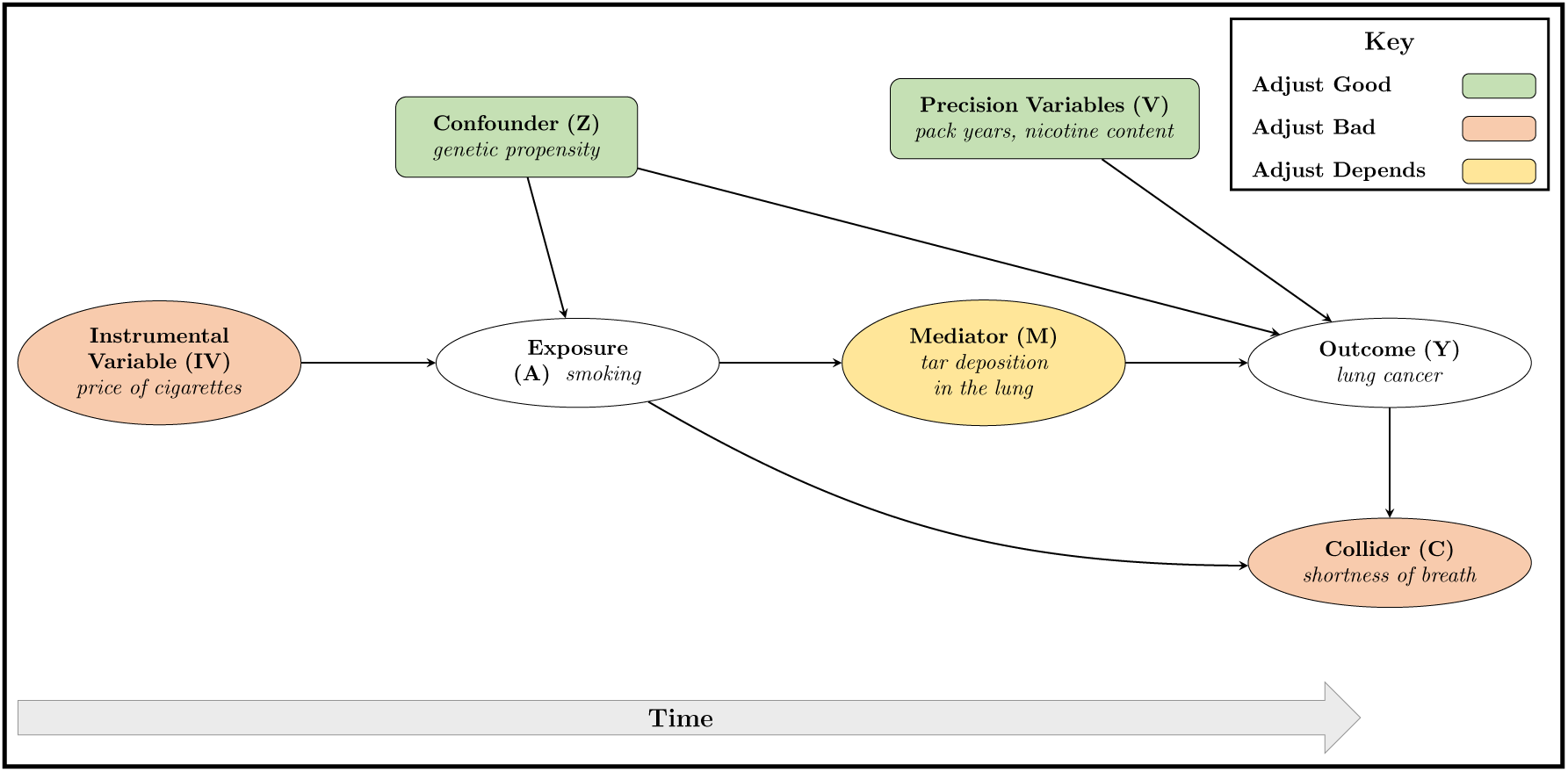
This figure depicts common third variable structures in the causal relationship between smoking (exposure) and lung cancer (outcome), represented as a directed acyclic graph (DAG) with temporal ordering along the x-axis. In a DAG, nodes denote variables and directed edges encode causal relationships. Third variables are factors related to the exposure and or outcome that are not the primary causal target. Confounders are common causes of both exposure and outcome that induce noncausal backdoor paths; conditioning on these variables blocks backdoor paths, also known as d-separation, and reduces bias. Precision variables are causes of the outcome that are not associated with the exposure; adjusting for them does not affect bias but can improve statistical efficiency. Instrumental variables are causes of the exposure that affect the outcome only through the exposure; conditioning on them can amplify bias in the presence of unmeasured confounding. Colliders are common effects of two variables, for example exposure and outcome; conditioning on a collider opens a noncausal path and induces spurious association, often referred to as collider stratification bias. Mediators lie on the directed causal path from exposure to outcome; conditioning on mediators blocks part of the total causal effect and yields estimates of the controlled direct effect rather than the total effect.

Directed acyclic graphs (DAGs), which represent variables as nodes and causal relations as directed edges, and structured confounder selection criteria, such as the disjunctive cause criterion/citevanderweele2019principles, have been proposed to guide variable selection and reduce subjective or ad hoc modeling decisions. Nonetheless, empirical assessments of published epidemiologic studies consistently document heterogeneity and inconsistency in the identification and handling of confounders, even for the same exposure–outcome relationships[10, 8, 9]. Figure 1 illustrates the role of a confounder in relation to an exposure and outcome of interest and contrasts this with other common third variable structures, including precision variables, colliders, instrumental variables, and mediators, each with distinct implications for adjustment and causal effect estimation.

To address these challenges, we propose an automated framework that uses large language models (LLMs) to support study screening and structured data extraction for systematic reviews and meta-analyses. The objective is to improve the efficiency and reproducibility of identifying eligible studies and extracting key study attributes, including exposures, outcomes, and candidate confounders. We introduce **LitScreen** and **VarEx**, two complementary prototype pipelines, implemented in Python and publicly available on GitHub, that apply LLM-based methods to prioritize and screen full text articles for inclusion and to extract and normalize reported study variables from observational research, without imposing additional causal modeling assumptions. Together, these tools aim to reduce manual burden while enabling more consistent and scalable characterization of the covariate sets and adjustment strategies actually used in published studies within evidence synthesis workflows. Finally, we map VarEx outputs into a structured representation compatible with the Metaconfoundr framework,[11] as operationalized in the Confounder Matrix, which groups covariates into higher level constructs, for example sociodemographic variables such as sex and age, genetic biomarkers such as APOe4, and health related behaviors, such as cigarette smoking, to support visualization and evaluation of construct level covariate coverage across studies.

### 1.1 Background

Given the ethical and practical constraints of randomized controlled trials for chronic conditions,[12] observational studies are crucial for understanding complex chronic diseases with multifactorial etiologies such as Alzheimer disease and related dementias (ADRD). However, these studies often exhibit substantial variability in confounder selection and adjustment. For example, prospective cohort studies are well suited to capturing health behaviors and family history but may be susceptible to recall bias, whereas retrospective studies are strong for ascertaining comorbidities but often lack detailed information on lifestyle factors.

This variability can lead to omitted variable bias, particularly in studies that lack data on key determinants such as APOE e2/e3/e4 genotype, repeated cognitive assessments, or neuroimaging. In addition, ambiguity in variable definitions, temporal ordering, and the assumed causal structure further complicates confounder control[13]. Even when investigators follow standardized recommendations, contradictory findings across studies may arise from underadjustment, overadjustment, or adjustment for inappropriate variables.

The first step in any research endeavor is to identify a corpus of relevant literature. This screening step is itself resource-intensive. Traditional systematic review methods require multiple reviewers to screen each citation manually against predefined inclusion criteria. Existing machine learning tools, such as Abstrackr, can reduce screening workload but often rely on proprietary platforms, which limits transparency and reproducibility[14, 15]. Recent advances in open source LLMs create new opportunities for transparent and reproducible automation of screening and data extraction workflows[16].

Our previous work developed methods for causal feature selection[17, 18, 19, 20] that operationalize causal criteria for third factor variables such as confounders by translating them into discovery patterns over structured knowledge sources derived from the literature and biomedical ontology grounded knowledge graphs[21]. These approaches show promise for identifying more comprehensive and systematically justified collections of candidate covariates for a given exposure–outcome relation, beyond what is typically reported in any single study. However, the absence of a scalable framework for jointly considering covariates reported in primary observational studies and variables suggested by graph theoretic discovery patterns constrains formal comparison and adjudication of these collections. This limitation impedes systematic identification of contradictions and potentially problematic adjustment variables at the design stage and hinders efforts to interpret robustness metrics such as E-values in terms of specific omitted or misspecified covariates and to develop field specific knowledge resources on expected confounders, as recommended in recent empirical work on E-value use[22, 23].

We will evaluate LitScreen and VarEx across multiple open datasets and targeted use cases in observational epidemiology. LitScreen will be developed and assessed using the SYNERGY dataset and in-house expert-curated reference standards, enabling rigorous evaluation of screening performance and error modes. VarEx will be applied to selected exposure-outcome pairs and evaluated against a single expert-constructed gold-standard extraction set to assess extraction accuracy and generalizability.

LLMs are large neural network models trained on massive text corpora to predict the next token in a sequence. Their capacity to process and generate natural language allows them to perform tasks such as classification, extraction, and summarization without task specific supervised training data. In contrast to rule based or traditional machine learning approaches that depend on hand-crafted rules or large labeled training sets, LLMs can leverage pre-trained knowledge and contextual understanding to extract nuanced information from complex textual data[24, 25].

A well recognized limitation of LLMs is hallucination, defined as the generation of content that is syntactically plausible but factually incorrect or unsupported by the input data[26]. To mitigate this risk, we propose a framework that incorporates retrieval augmented generation, human oversight, and explicit evaluation against independent reference standards. Retrieval augmented generation combines LLMs with document retrieval so that the model conditions its responses on relevant source texts rather than relying solely on internal parameters[27]. In our framework, LLM outputs for screening and extraction are systematically compared with human annotated gold standards to quantify accuracy and identify failure modes.

The proposed framework includes three main components: (1) retrieval augmented prompting with domain specific instructions to guide the LLM, (2) a multi stage automated extraction and classification process, and (3) evaluation against an independent human annotated reference standard to assess the accuracy and completeness of extracted data. The goal is to assess the feasibility and potential benefits of using LLMs to assist study screening and variable extraction in epidemiologic research, while explicitly addressing hallucination and other sources of error.

The objectives of this research developing and evaluating two complementary automated pipelines for screening studies and extracting key study variables are enumerated below. Specifically, we aim to:

1. **Develop an automated screening pipeline.** Design and implement **LitScreen**, a prototype pipeline that uses large language models (LLMs) to automate title and abstract screening for systematic reviews by applying configurable inclusion criteria to identify eligible observational studies.
2. **Develop an automated variable extraction pipeline.** Implement **VarEx**, a prototype pipeline that uses LLMs to extract exposures, outcomes, and covariates from full text articles, and establish a benchmark for identifying and extracting essential study variables in studies of hypertension as a risk factor for ADRD.
3. **Classify extracted covariates.** Assign each extracted covariate to a predefined semantic category (for example, sociodemographic, clinical, behavioral, or genetic), generating structured, machine readable output suitable for downstream analyses, including Metaconfoundr based assessment of confounder coverage.
4. **Database of Covariates Reported in the Published Literature.** LitScreen and VarEx outputs can be used to create a structured, reusable field specific covariate databases cataloging exposures, outcomes, and covariates reported in published observational studies. These covariates can then be compared with those suggested by graph theoretic discovery over biomedical knowledge graphs in order to inform future causal feature selection and evaluation of confounder coverage.
5. **Evaluate performance and generalizability.** Quantify screening and extraction performance using standard metrics, including sensitivity, precision, recall, and F_1_, and assess whether the pipelines generalize to exposure–outcome relationships beyond hypertension and ADRD.
6. **Quantify efficiency gains.** Compare per document processing time for LitScreen and VarEx with manual review baselines to estimate time savings across the screening and extraction stages of systematic reviews.

Through these objectives, we seek to provide a scalable, reproducible approach for managing and analyzing complex observational datasets and for characterizing how covariate adjustment is implemented in practice across epidemiologic studies.

## 2 METHODS AND MATERIALS

In this study, we developed an end-to-end workflow comprising two complementary pipelines that leverage large language models (LLMs). LitScreen automates the screening of observational studies, and VarEx automates the extraction of their study variables. As shown in Figure 2, the process begins with a health sciences librarian developing a reproducible MEDLINE search via OVID, which generates an RIS file. Because RIS files consist of both relevant and irrelevant papers, irrelevant papers are first screened out by LitScreen, a three-phase LLM screening stage, retaining only those that satisfy predefined inclusion criteria. After filtering out irrelevant papers, we extract nuanced information by feeding the relevant papers to the VarEx variable extraction stage.

**Figure 2:**
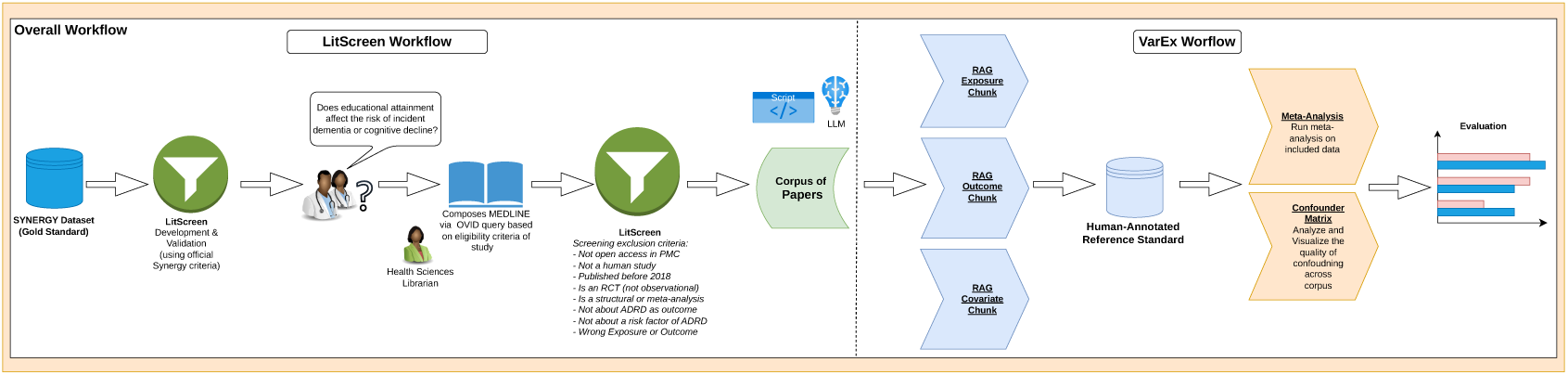
End-to-end workflow of LitScreen and VarEx. LitScreen screens papers retrieved via MEDLINE, and VarEx extracts exposure, outcome, and covariate evidence through a RAG-augmented LLM, with extraction performance evaluated using confusion matrices.

The LitScreen pipeline decomposes the screening task into three steps: abstract-level sentence extraction, abstract-level decision-making based on the screening criteria, and full-text RAG-based verification. We progressively add more evidence to fewer papers to reduce computational cost, achieving up to 90% savings in some systematic reviews. After deduplication, all records are sent for abstract-level screening, where highly irrelevant papers are excluded; the remaining small portion of papers is sent for full-text retrieval-augmented verification, where relevant sections are extracted and submitted for a second round of LLM-based adjudication. This staged design applies the most computationally expensive processing only to the smallest and most uncertain subset of records.

The papers retained after screening are then passed to the variable extraction stage, which uses LLMs to extract exposures, outcomes, and covariates from their full text. Throughout the text, we point out where additional details are available in the appendices located in the supplementary material or this paper’s accompanying Zenodo database. GitHub was used for version control and collaborative work, and is free and publicly available.

### 2.1 Data Sources and Corpora

We will evaluate the use and performance of LitScreen and VarEx across multiple open datasets and application scenarios.

For LitScreen, we will use the SYNERGY dataset for model development and evaluation. SYNERGY includes ground truth screening labels for 169,288 publications drawn from 26 systematic reviews, of which 2,834 (1.67%) were deemed eligible. We will focus on a subset of five systematic reviews addressing observational epidemiologic questions, comprising 52,541 publications. We used labeled reference datasets from the SYNERGY benchmark [28] to evaluate the performance of our pipeline because the data come from publicly available, free systematic reviews, previously screened and labeled by human experts. Since all the citations used in the dataset have been screened by human experts, it serves as a gold standard, allowing us to validate the classification accuracy and reliability of our pipeline against established expert decisions. The SYNERGY benchmark provides ground-truth inclusion labels assigned by two independent reviewers, with disagreements resolved by consensus. We used six datasets whose inclusion criteria explicitly require observational or longitudinal human study designs, consistent with our target use case (Table 1).

**Table 1:**
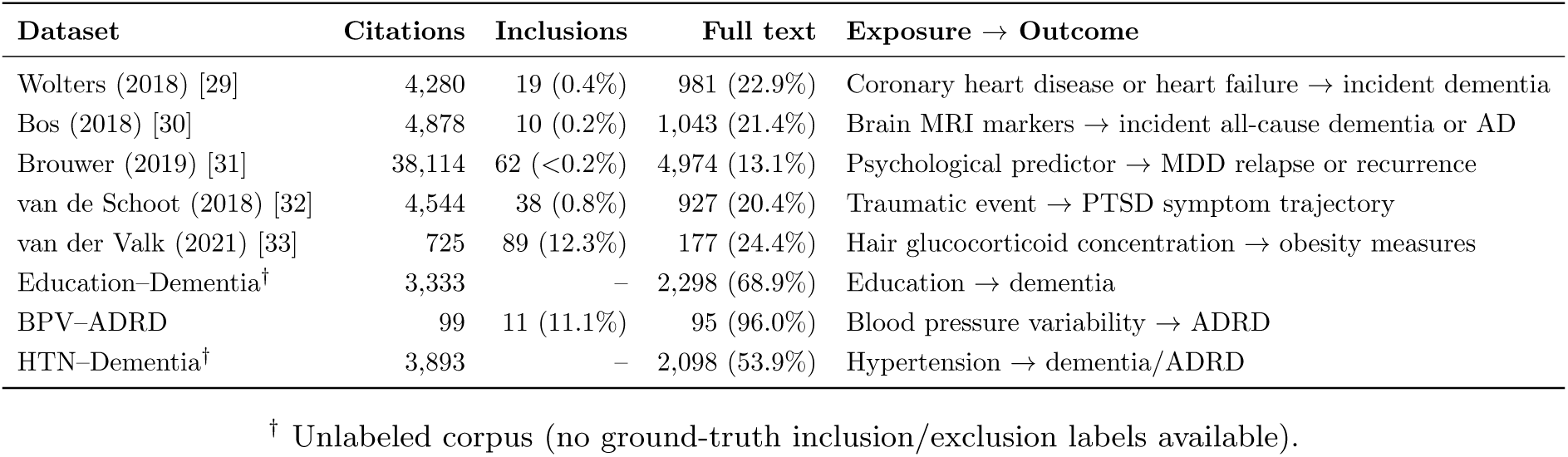
SYNERGY benchmark datasets and production corpora used for screening validation.

#### MONKEY MONKEY MONKEY

TO ADD TO DO DO ADD TO DO TO ADD TO DO TO ADD TO DO To complement these data, we developed two in-house reference standards using subject matter experts. These include studies of blood pressure variability as a risk factor for Alzheimer’s disease and related dementias (ADRD), and studies evaluating PTSD treatments in relation to self-injury, self-harm, and suicidality outcomes. We selected these topics because the resulting literature sets were of manageable size for detailed screening evaluation, each yielding fewer than 200 records from our query, compared with the 3,000 to 6,000 records typical of systematic reviews. These reference standards enabled detailed error analysis and iterative refinement of screening performance.

For VarEx, we will evaluate three exposure-outcome pairs: PTSD and self-harm or suicidality, hypertension and ADRD, and education and ADRD. For each pair, subject matter experts will construct reference standards through manual extraction of study-level information.

#### MONKEY MONKEY MONKEY

Once we achieved acceptable performance, we applied the LitScreen pipeline to additional use cases and used its outputs as inputs (corpora) to the VarEx pipeline.

We also evaluated our pipeline on unlabeled datasets of large and diverse collections of citations: a dataset examining education (exposure) as a risk factor for dementia (outcome), and another examining hypertension (exposure) as a risk factor for Alzheimer’s Disease and Related Dementias (ADRD) or Mild Cognitive Impairment (MCI) (outcome). These are unlabeled datasets exported directly from bibliographic databases in the Research Information System (with the “RIS” file-name extension) and lacked ground-truth labels. The performance on these datasets was evaluated manually; human experts reviewed the final subset of papers selected by the LLM to verify whether they met the defined eligibility criteria and the required study designs.

Our search for these production datasets was first devised as a structured, reproducible MEDLINE search via OVID queries, in collaboration with a health sciences librarian. We used MEDLINE via Ovid because its controlled indexing and consistent search functionality support reproducible literature retrieval. We used the RIS outputs from our MEDLINE via OVID queries as the starting point for our corpus. These queries are available in the supplementary material in Appendix 6. Also, to promote reproducibility and due to copyright concerns, we used only open-access full-text articles available via PubMed Central or open-access versions identified via Unpaywall.

The primary datasets for this study consisted of observational studies examining hypertension (HTN) as an exposure or risk factor for dementia or ADRD. This HTN*→*ADRD corpus represents the main scientific use case for VarEx and was used for the primary variable extraction and evaluation analyses. To evaluate whether the extraction pipeline generalized beyond the primary HTN*→*ADRD use case, we also applied the same extraction and classification workflow to additional observational-study corpora involving different exposure–outcome relationships. These secondary corpora were not the main scientific target of the study; rather, they served as validation datasets for assessing whether VarEx could identify exposures, outcomes, and adjusted covariates across epidemiologic studies outside the HTN*→*ADRD domain.

### 2.2 Screening Pipeline

We designed a three-phase screening module to simulate the cognitive processes of human expert reviewers and to enable efficient screening. Figure 3 outlines the overall screening workflow, proceeding from formulation of the clinical research question (Step 1), through the MEDLINE database search (Step 2), to structured eligibility screening (Step 3), which yields the corpus of papers. Instead of forcing a large language model (LLM) to read the full text and provide an inclusion/exclusion decision in a single prompt, we decomposed our screening process into multiple tasks. The process starts by extracting evidence from the abstract, and subsequently applies more complex reasoning and heavier computational resources only to the subset of citations that require them.

**Figure 3:**
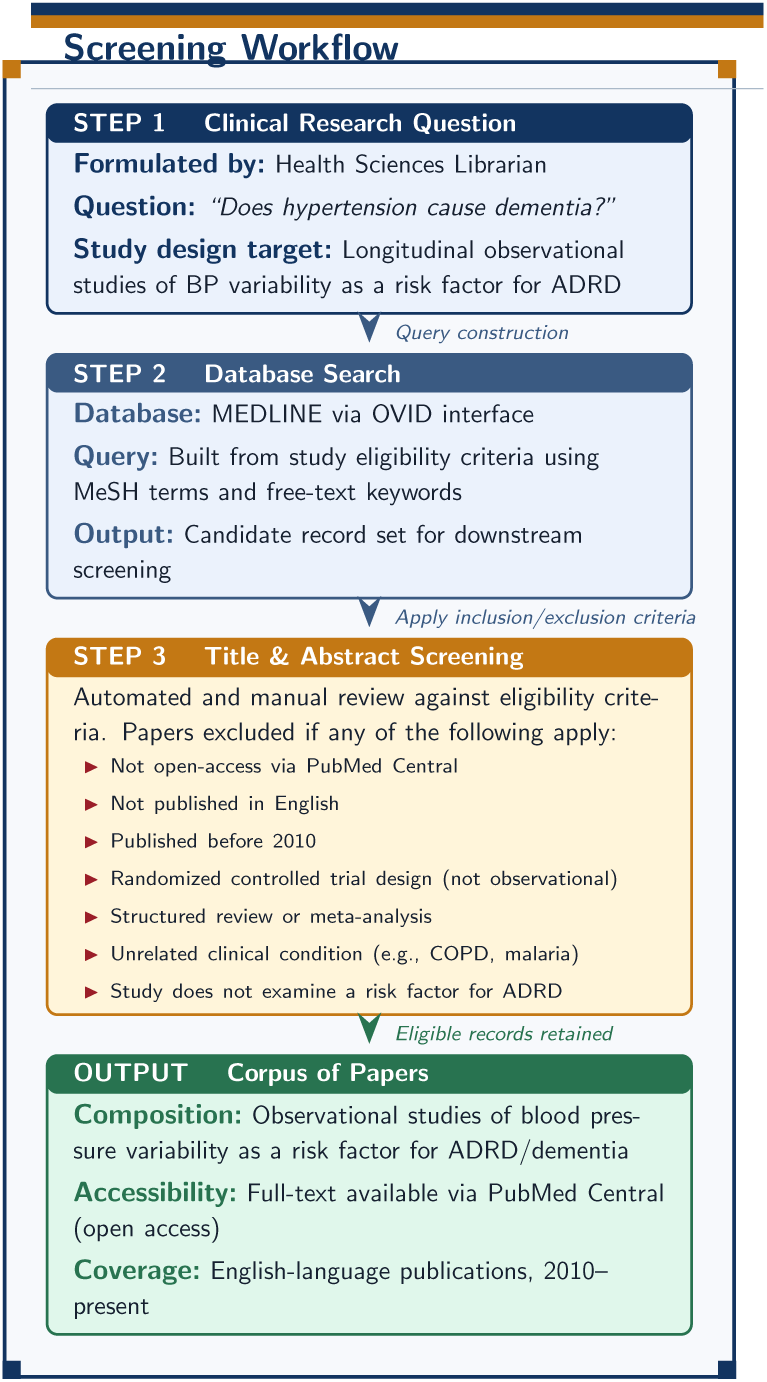
*Screening workflow for developing the corpus of papers.* The pipeline proceeds from clinical question formulation (Step 1) through MEDLINE database search (Step 2) to structured eligibility screening (Step 3), yielding a final corpus of open-access observational studies (2010–present) examining blood pressure variability as a risk factor for Alzheimer’s disease and related dementias (ADRD).

**Figure 4:**
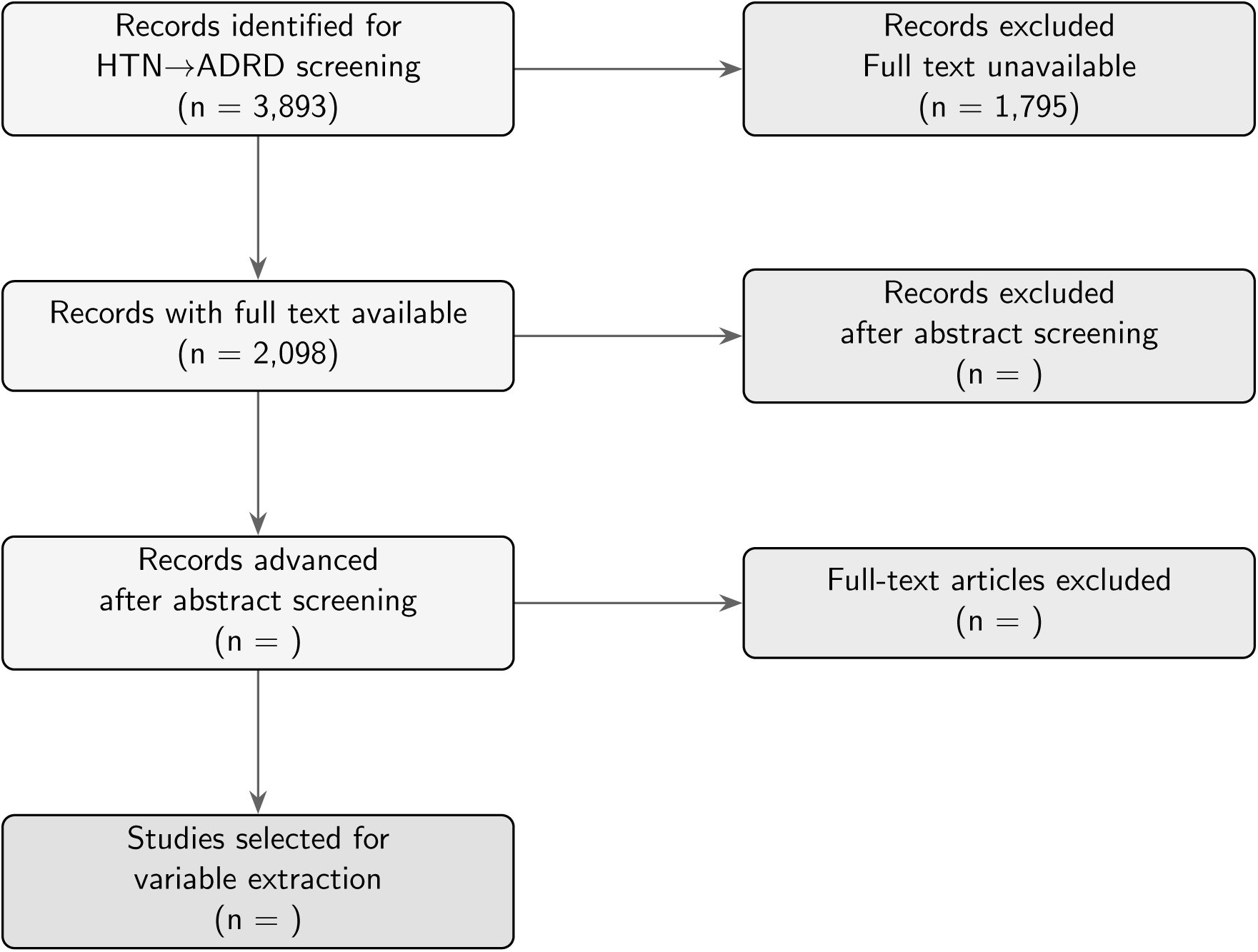
PRISMA [44] diagram of the screening process for our HTN*→*ADRD corpus, using abstract and full-text as input. Identification of open access (PubMed Central) human observational studies studying hypertension and dementia/ADRD. Records removed before abstract and full-text screening. Records screened for open access. Reports sought for retrieval. Reports assessed for eligibility. Studies included in corpus for variable extraction.

#### 2.2.1 Phase 1: Domain-Neutral Sentence Extraction

The first phase of the screening pipeline acts as a few-shot information extractor, for which we used llama3.3:70b. We engineered the prompt to be completely domain-neutral to prevent the model from introducing bias or making premature screening decisions. All specific names of exposures, outcomes, or study designs were deliberately removed from both the prompt instructions and the few-shot examples so that the model does not merely pattern-match keywords while extracting evidence sentences from the abstract. Instead, the prompt strictly instructs the model to extract only the sentences from the abstract that describe the study’s design, interventions/exposures, and measured outcomes. This approach forces the model to make decisions based on the extracted evidence rather than the entire abstract.

#### 2.2.2 Phase 2: Criterion-Wise Decision-Making on Extracted Evidence

In the second phase, we narrow down the candidate pool before any full-text retrieval is attempted, based solely on the verbatim sentences extracted from the abstract. To reduce the candidate pool of citations, we engineered an independent prompt for Deepseek-R1:70b, a reasoning-tuned model that emits its chain of thought within <THINK> tags before committing to a structured answer. The prompt enforces a three-step procedure: a preliminary analysis, a self-reflection analysis against a set of 4–5 criterion-specific questions, and a final categorical answer (YES/NO/UNCLEAR). Additionally, it includes a self-assessed level of confidence (HIGH/MEDIUM/LOW) and a typed evidence field that names the structural element identified (e.g., DESIGN TYPE = “prospective cohort”). The model does not make the final disposition of the paper; that is determined by a deterministic three-way gate. If the LLM answers NO for any criterion with high confidence, the paper is immediately excluded without further consideration. All other cases—an UNCLEAR answer, any non-HIGH confidence, or any disagreement among criteria—are routed to the full-text screening phase. This restricts the more expensive full-text computation to genuinely unresolved criteria and to papers recommended as Include during abstract screening.

#### 2.2.3 Phase 3: Full-Text Retrieval-Augmented Verification

Papers that are escalated by the abstract gate are sent to the retrieval-augmented verification phase, where the LLM reasons over passages drawn from the article rather than the abstract alone. This phase consists of four components: full-text acquisition, structure-aware parsing and chunking, two-stage retrieval, and final LLM verification.

##### Full-text acquisition

For each escalated paper, we extracted its PubMed ID (PMID) and PubMed Central ID (PMCID) from its DOI using the NCBI Entrez research endpoint, then retrieved the full text in JATS XML format. For papers without an open-access PMC record, we attempted to retrieve an open-access PDF via Unpaywall and converted it to JATS XML format. Articles for which neither source yields full text are flagged as MISSING FULLTEXT for manual review rather than auto-exclusion.

##### Structure-aware parsing and chunking

Each XML file is converted into a typed ParsedDocument using a custom JATS parser. This conversion allows us to classify header keywords and retain only the Methods and Results sections for downstream retrieval. All other sections, such as Introduction, Discussion, Acknowledgments, and References, are dropped because they tend to summarize the work of others and degrade retrieval precision. Each retained section is split into sentences using a scientific-text-aware regex (preserving decimals, common abbreviations, and inline citations) and grouped into semantic chunks using a greedy, adaptive procedure. The procedure starts with a seed sentence; subsequent sentences are appended to the current chunk while their embedding similarity to a running-average chunk embedding (weighted 0.7 toward the existing chunk and 0.3 toward the newly added sentence) exceeds 0.80, and the chunk remains below 500 words. If any chunk has fewer than 60 words, it is reattached to its predecessor. The output is a per-section sequence of semantically coherent passages labeled Section – Chunk N.

##### Two-stage retrieval

We wrote a multi-sentence query phrased to match the language of Methods/Results content for each criterion (Appendix 6). All queries and chunks were embedded with bioLinkBERT-large, a biomedical-domain transformer pretrained on PubMed abstracts and citation links. The first retrieval stage used cosine similarity to score text chunks, then applied Maximal Marginal Relevance (*λ* = 0.7) to select the top 50 passages across sections, balancing relevance and redundancy. In the second stage, these 50 candidates are rescored with the BAAI/bge-reranker-large cross-encoder, which jointly attends over the query and the chunk text, and retains only the top 5 for the LLM. The two-stage design restricts the expensive cross-encoder pass to a small candidate pool while preserving recall through the cheaper first stage.

##### LLM verification

The five retrieved chunks for each unresolved criterion were assembled into a context block, prefixed with the verbatim abstract sentences originally extracted in Phase 1 (labeled as such). These were then submitted to Deepseek-R1:70b under a verification prompt structurally analogous to Phase 2, but with two key differences: the model is not allowed to return UNCLEAR (since the evidence base is now significantly richer than an abstract), and the prompt instructs it to cite the chunk identifier that justifies its answer. Finally, paper-level inclusion is computed by combining the high-confidence YES criteria from the abstract phase with the answers carried forward from full-text verification under the same criteria: any NO excludes, and all YES includes.

#### 2.2.4 Prompt Engineering for Screening

Screening prompt design was driven by a single overriding objective: maximize recall — that is, ensure no eligible paper is discarded — while still reducing the pool of papers that require manual review. This asymmetric cost structure (a missed eligible paper is far worse than a borderline inclusion) shaped every design decision described below.

##### Criteria sourcing

The eligibility criteria fed into the prompts were not standardized across datasets. For the six SYNERGY benchmark datasets, we used the official inclusion and exclusion criteria published in the original systematic review protocol. For the three production datasets (Hypertension*→*ADRD, Education*→*Dementia, BPV*→*Dementia), domain experts provided the criteria they themselves applied during manual screening. In both cases, criteria were operationalized at the *criterion level* : each individual criterion was instantiated as its own prompt, independent of the total number of criteria in a given review. A review with three criteria simply invokes the same template three times.

##### Evidence-basis gate

All decision prompts — both abstract-level (Phase 2) and full-text (Phase 3) — share a mandatory evidence classification step that the model must complete before selecting an answer. The model must declare one of three evidence states: Disqualifying (an extracted sentence affirmatively states that the paper violates the criterion and can be quoted verbatim), Confirming (an extracted sentence affirmatively states the criterion is met), or Insufficient (no extracted sentence directly settles the question). This declaration mechanically constrains the final answer: a No answer is only permitted when the evidence state is Disqualifying. At the abstract stage, Insufficient evidence maps to Unclear, which routes the paper to full-text screening. At the full-text stage, Insufficient maps to No — because there is no further stage to escalate to, and excluding a paper without a verbatim disqualifying quote is not permitted.

##### Three-stage reasoning structure

Both Phase 2 and Phase 3 prompts enforce a three-stage structured chain of thought: an initial analysis (Stage 1), a self-reflection pass that critiques the initial answer against a set of 4–5 criterion-specific reflection questions (Stage 2), and a final answer (Stage 3). This structure was chosen to reduce impulsive NO decisions that frequently arise when a model commits to an answer before considering confusable cases. The reflection questions are tailored to each criterion and encode the most common over-reads observed during development — for example, distinguishing a cross-sectional *analysis* from a cross-sectional *design*, or distinguishing non-systemic hypertension subtypes (gestational, portal) from systemic hypertension.

##### Deterministic verbatim gate

To prevent the model from manufacturing a disqualifier that is not present in the evidence, Phase 2 prompts require the model to populate a DISQUALIFIER VERBATIM output field with an exact, word-for-word copy of the sentence that justifies any No answer. A downstream deterministic code gate then verifies that this quote is a contiguous verbatim substring of the extracted sentences. If the quote is a placeholder (“N/A”), paraphrased, or cannot be located in the input, the No is automatically demoted to Unclear and the paper proceeds to full-text screening.

Examples of the abstract evidence extraction prompt are provided in Appendix 6, and the abstract decision prompt and full-text decision prompt are provided in Appendix 6.

### 2.3 Variable Extraction Pipeline

We developed a retrieval-augmented large language model pipeline to extract epidemiological study variables from all relevant and open-access full-text biomedical articles. The pipeline extracted three types of variables (exposure, outcome, and covariates) from the abstract, Methods, and Results sections of these articles. First, we parsed the XML structure to select relevant sections, divided them into semantically coherent chunks, retrieved role-specific evidence, and then used a large language model to extract variables along with supporting evidence. All extracted covariates were assigned to predefined semantic categories in a separate classification step.

#### 2.3.1 Section Selection and Retrieval

Full-text articles were processed in PMC/JATS/XML format using the same structure-aware parser and adaptive chunking procedure described for the screening pipeline’s full-text verification phase (Section 2.2.3). Unlike the screening stage, which retained only the Methods and Results sections, the extraction stage additionally retained the abstract. This restriction to the abstract, Methods, and Results sections was guided by Strengthening the Reporting of Observational Studies in Epidemiology (STROBE), which recommends reporting key study design information in the abstract, detailed variable definitions and statistical methods in the Methods section, and participant characteristics, outcome data, and the main analyses in the Results section [34]. In addition, the pipeline extracted tables, their captions, and figure captions (excluding the figures themselves), structured as text chunks so that information reported in table captions, contents, and footnotes is included in retrieval.

Chunks were retrieved in the same two-stage manner as the screening pipeline, with two differences. First, chunks and queries were embedded with NeuML/pubmedbert-base-embeddings [35], a PubMedBERT-based [36] sentence-transformer fine-tuned on PubMed title–abstract pairs to produce dense embeddings for biomedical semantic search. Second, exposure, outcome, and covariate were retrieved separately using role-specific query templates (Appendix 6) designed to capture exposure definitions, outcome definitions, and statistical adjustment language. Candidate chunks were first ranked by embedding similarity and then re-ranked with the BAAI/bgereranker-large cross-encoder [37], after which the pipeline supplied up to four chunks per role to the language model for extraction.

#### 2.3.2 Prompt Engineering

VarEx uses a structured prompt-engineering strategy. We used separate prompts for exposures, outcome, and covariates to ensure the model extracts one variable role at a time. This modular decomposition of a complex extraction task into simpler, single-role subtasks follows established prompting practice [38], and is intended to reduce common extraction errors such as misclassifying outcomes as exposures, treating baseline characteristics as covariates, or summarizing the study rather than listing variables.

In order to design an effective prompt, we took inspiration from widely accepted chain-of-thought-style [39] task decomposition, but the pipeline did not require the model to output an unrestricted reasoning trace. Instead, the prompts used explicit definitions, stepwise instructions, decision rules, and checklist-style constraints to guide the model toward the final structured output. Exposures were defined as independent variables, risk factors, or predictors under study; outcomes were defined as dependent variables, endpoints, or diagnoses under study, or the measured outcome of the study; and covariates were defined as variables that were statistically controlled for or adjusted for in regression or multivariable models.

To improve evidence grounding, each extracted variable had to contain an exact supporting quotation taken from the retrieved chunks, together with the chunk identifier. Because the model tended to generate statistical conclusions and p-values, we added a statement to the prompt instructing it not to generate any statistical conclusions. For covariate extraction, the model was instructed to list each adjusted variable separately when multiple variables appeared in the same adjustment statement. The role outputs were collected into role-specific lists, and the supporting quotations were stored separately for auditability and manual review.

#### 2.3.3 Models and Configuration

Table 2 summarizes the models and key configuration parameters used in the extraction pipeline. All parameters were held constant across articles to ensure consistency. We report results using the open-source Deepseek-R1:70b and Llama3.3:70b models, but the pipeline is provider-agnostic and is equally compatible with paid commercial models such as OpenAI’s GPT-4o [40].

**Table 2:** Models and configuration parameters used in the screening and extraction pipelines.

| Component | Setting |
| --- | --- |
| *LitScreen Models* |  |
| Embedding model (bi-encoder retrieval) | bioLinkBERT-large [41] |
| Cross-encoder reranker | BAAI/bge-reranker-large |
| Relevance/Extraction LLM (Phase 1) | llama3.3:70b [16] |
| Decision LLM (Phase 2 & 3) | Deepseek-R1:70b[42] |
| *VarEx Models* |  |
| Embedding model (bi-encoder retrieval) | NeuML/pubmedbert-base-embeddings |
| Cross-encoder reranker | BAAI/bge-reranker-large |
| Extraction LLM | Deepseek-R1:70b [42] (open-source, via Ollama), temperature = 0.3 |
| *VarEx Retrieval and Chunking Parameters* |  |
| Chunk similarity threshold | 0.80 (cosine) |
| Chunk size | 60–500 words |
| Candidate chunks before reranking | up to 20 |
| Final chunks retrieved per role | up to 4 |

#### 2.3.4 Covariate Classification

At this stage of the pipeline, we have extracted covariates from the relevant biomedical articles. In the next step, these extracted covariates are classified using the same language model. Each covariate was assigned to exactly one of ten predefined semantic categories:

1. Sociodemographics
2. Biology/Genetics
3. Health Behaviors
4. Medication Use/Drug Exposure
5. Physiological Measurement
6. Psychosocial/Life Events
7. Clinical Symptoms/Severity Measurement
8. Psychiatric/Medical History
9. Underlying Health Conditions
10. Misc/Other

The classification prompt included category definitions, examples, and disambiguation rules. These rules addressed common sources of ambiguity, such as the distinction between current chronic conditions and prior medical history, the distinction between physiological measurements and diagnosed conditions, and the classification of clinical severity scores.

### 2.4 Evaluation

The evaluation focuses on assessing the efficacy and reliability of integrating large language models (LLMs) into systematic reviews and meta-analyses. We evaluated the screening pipeline and the variable extraction pipeline separately. For screening, we assessed how accurately the pipeline reproduced expert inclusion/exclusion decisions; for extraction, we assessed the precision and consistency of the extracted variables against an independent, human-annotated reference standard.

#### 2.4.1 Screening Evaluation

We validated our screening pipeline using six systematic review datasets from the SYNERGY benchmark [28] and applied it prospectively to two unlabeled production datasets. Because all citations in the SYNERGY datasets were previously screened by human experts, these labels served as a gold standard against which we validated the classification accuracy and reliability of our pipeline. For the two unlabeled production datasets (education and hypertension), performance was evaluated manually; human experts reviewed the final subset of papers selected by the LLM to verify whether they met the defined eligibility criteria and the required study designs.

The search queries were intentionally restrictive, reducing the number of studies to screen and minimizing the burden on the manual reviewers. To compare the automated screener against manual expert review, a sample size of approximately 100 papers was selected for its statistical soundness and feasibility. This sample size is sufficient to detect moderate-to-large differences in screening accuracy metrics, such as sensitivity and specificity, which we quantified using Cohen’s *h*, a measure of effect size. With 80% power and a two-sided test at a significance level (*α*) of 0.05, a sample of 100 can detect a Cohen’s *h* of 0.4 or greater, indicating moderate-to-large differences in accuracy. This power threshold aligns with standard biostatistical practice for screening studies.

#### 2.4.2 Extraction Reference Standard

We constructed a manually annotated reference standard for evaluation of extraction and classification. Each paper was reviewed independently by human experts, who recorded exposures, outcomes, and covariates, and assigned each covariate to one of the ten predefined semantic categories. The Hypertension*→*ADRD corpus was treated as the primary reference-standard dataset for evaluating VarEx, while additional observational-study corpora involving other exposure–outcome relationships were treated as external validation datasets. All datasets were evaluated using the same extraction, semantic matching, and covariate classification metrics, allowing us to assess both performance on the main dementia-focused research question and generalizability across different epidemiologic domains.

#### 2.4.3 Variable Extraction and Classification Performance

The reference standard and extracted variables were semantically matched to evaluate extraction performance. Each variable name was embedded with NeuML/pubmedbert-base-embeddings, and cosine similarity was computed between every reference and extracted variable. Variables were matched one-to-one using the Hungarian algorithm [43], ensuring that each reference and each extracted variable was assigned to at most one counterpart. This one-to-one constraint prevents a single variable from being double-matched against two reference covariates. Each pair of matched variables was accepted as a true positive only if cosine similarity met or exceeded a threshold of 0.60. A sensitivity analysis across threshold values, and the rationale for selecting 0.60, is reported in Appendix 6.

We assessed performance in two stages. In the extraction stage, all reference and extracted covariates were compared as flat lists, ignoring category labels, to measure how well the pipeline identified the correct variables. From the resulting true positives, false positives, and false negatives, we computed precision, recall, and F1. In the classification stage, we assessed whether correctly extracted covariates were also assigned to the correct semantic category. We report:

- classification accuracy, defined as the proportion of correctly matched covariates assigned to the correct category;
- per-category precision, recall, and F1, together with the macro-averaged F1 across categories;
- a strict combined score that counts a covariate as correct only when both the variable and its category match the reference standard.

The difference between the extraction F1 and the strict combined F1 quantifies the performance lost to misclassification.

## 3 RESULTS

The results of our study demonstrate the feasibility and potential benefits of using LLMs to facilitate screening and data extraction in systematic reviews and meta-analyses. LitScreen effectively filtered relevant papers from extensive databases, focusing on human studies related to dementia and ADRD while excluding meta-analyses and non-observational studies. This process ensured that the literature to be reviewed was relevant and focused.

The results using the full text are included in Appendix 6 of the Supplementary Material. The data extraction phase, which used LLMs to process and analyze the selected papers, yielded comprehensive and accurate datasets. The LLMs successfully identified, extracted, and organized detailed information from complex texts, focusing on predefined categories such as study design, participant demographics, outcomes measured, and confounders controlled for. The multi-stage extraction process, which included initial extraction, refinement and validation, contextual analysis, consolidation, and categorization, ensured the accuracy and relevance of the extracted data.

### 3.1 Extraction Performance of VarEx

Table 3 shows the performance of VarEx in extracting variables for the primary Hypertension*→*ADRD reference standard and a secondary reference-standard dataset used for additional validation. We used studies focused on hypertension as a risk factor for ADRD and treated the Hypertension*→*ADRD as the primary evaluation dataset.

**Table 3:**
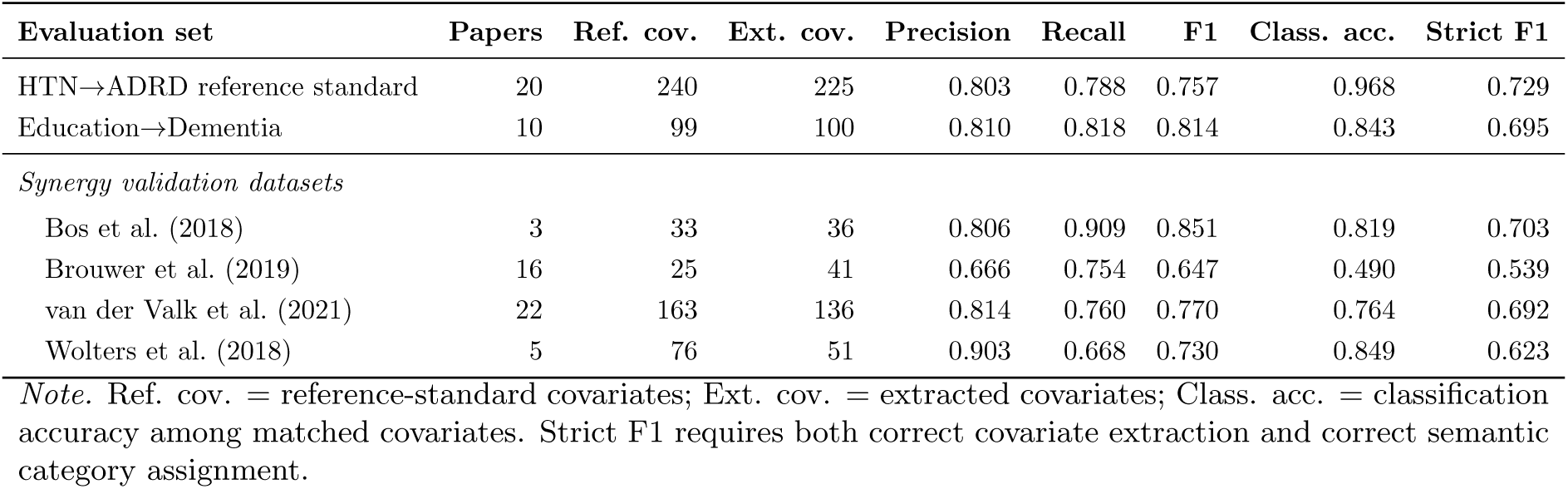
Overall extraction and classification performance across reference-standard evaluation sets.

| Evaluation set | Papers | Ref. cov. | Ext. cov. | Precision | Recall | F1 | Class. acc. | Strict F1 |
| --- | --- | --- | --- | --- | --- | --- | --- | --- |
| HTN→ADRD reference standard | 20 | 240 | 225 | 0.803 | 0.788 | 0.757 | 0.968 | 0.729 |
| Education→Dementia | 10 | 99 | 100 | 0.810 | 0.818 | 0.814 | 0.843 | 0.695 |
| *Synergy validation datasets* |  |  |  |  |  |  |  |  |
| Bos et al. (2018) | 3 | 33 | 36 | 0.806 | 0.909 | 0.851 | 0.819 | 0.703 |
| Brouwer et al. (2019) | 16 | 25 | 41 | 0.666 | 0.754 | 0.647 | 0.490 | 0.539 |
| van der Valk et al. (2021) | 22 | 163 | 136 | 0.814 | 0.760 | 0.770 | 0.764 | 0.692 |
| Wolters et al. (2018) | 5 | 76 | 51 | 0.903 | 0.668 | 0.730 | 0.849 | 0.623 |

*Note.* Ref. cov. = reference-standard covariates; Ext. cov. = extracted covariates; Class. acc. = classification accuracy among matched covariates. Strict F1 requires both correct covariate extraction and correct semantic category assignment.

In addition to overall extraction performance, we also analyzed the performance across covariate types. In Table 4, we have shown the category-level performance across all the biomedical articles which passed the screening criteria. The pipeline performed best for categories with more frequent and clearly stated covariates, including Health Behaviors, Sociodemographics, Medication Use/Drug Exposure, and Physiological Measurement. In contrast, performance was lower for sparse or semantically ambiguous categories, particularly Psychiatric/Medical History and Misc/Other. These categories contained few reference-standard covariates and often overlapped conceptually with other categories, making classification more difficult. For example, medical history terms may be confused with Underlying Health Conditions when the text does not clearly distinguish prior events from current baseline comorbidities, while Misc/Other is a heterogeneous residual category with less consistent linguistic patterns.

**Table 4:** Category-level covariate performance in the HTN*→*ADRD reference standard.

| Covariate category | Ref. cov. | Ext. cov. | Precision | Recall | F1 |
| --- | --- | --- | --- | --- | --- |
| Biology/Genetics | 5 | 4 | 1.000 | 0.800 | 0.889 |
| Health Behaviors | 42 | 41 | 0.878 | 0.857 | 0.867 |
| Sociodemographics | 77 | 65 | 0.954 | 0.805 | 0.873 |
| Medication Use/Drug Exposure | 17 | 11 | 0.727 | 0.471 | 0.571 |
| Physiological Measurement | 20 | 19 | 0.895 | 0.850 | 0.872 |
| Psychosocial/Life Events | 3 | 3 | 0.333 | 0.333 | 0.333 |
| Clinical Symptoms/Severity Measurement | 9 | 3 | 1.000 | 0.333 | 0.500 |
| Psychiatric/Medical History | 13 | 11 | 0.727 | 0.615 | 0.667 |
| Underlying Health Conditions | 48 | 62 | 0.597 | 0.771 | 0.673 |
| Misc/Other | 6 | 6 | 0.833 | 0.833 | 0.833 |

*Note.* Ref. cov. = reference-standard covariates; Ext. cov. = extracted covariates. Category-level performance requires both covariate identification and assignment to the correct semantic category.

### 3.2 Covariate Classification Performance

Our covariate classification performance classified 91 number of covariates for biomedical article focused on HTN and ADRD and 434 number of covariates for biomedical articles focused on other exposure and outcome. Table 5 summarizes classification performance across these two reference-standard datasets, including the number of matched covariates, classification accuracy, precision, recall, and F1 score.

**Table 5:** Classification performance: accuracy, precision, recall, and F1 score for correctly extracted covariates assigned to one of ten covariate categories.

| Dataset | Matched cov. | Class. acc. | Recall | Precision | F1 |
| --- | --- | --- | --- | --- | --- |
| HTN→ADRD | 189 | 0.958 | 0.667 | 0.794 | 0.708 |
| BPV→ADRD | 91 | 0.901 | 0.800 | 0.785 | 0.786 |
| Education→Dementia | 83 | 0.843 | 0.710 | 0.622 | 0.636 |
| *Synergy validation datasets* |  |  |  |  |  |
| Bos et al. (2018) | 30 | 0.819 | 0.751 | 0.665 | 0.703 |
| Brouwer et al. (2019) | 17 | 0.490 | 0.567 | 0.575 | 0.539 |
| van der Valk et al. (2021) | 122 | 0.764 | 0.687 | 0.727 | 0.692 |
| Wolters et al. (2018) | 47 | 0.849 | 0.572 | 0.768 | 0.623 |

*Note.* Matched cov. = covariates correctly extracted in Stage 1 (true positives), the basis for classification evaluation; Class. acc. = classification accuracy among matched covariates. Recall, precision, and F1 are macro-averaged across the ten covariate categories, treating category assignment as a multi-class classification problem evaluated only on matched covariates.

### 3.3 Efficiency Analysis: Manual Review Time Versus LLM-Assisted Pipeline Runtime

Improvements in time efficiency were evaluated by breaking the screening and extraction workflow into three stages: i) abstract screening, ii) full-text screening, and iii) data extraction. As reported by Wang et al., average processing times per reference per reviewer were 0.9 minutes for abstract screening, 7 minutes for full-text screening, and 53 minutes for data extraction [45, 46]. Table 6 represents the per-document processing time comparison at each stage. Overall, the pipeline saves 85.7% or approximately 52 minutes of time per reference.

**Table 6:** Processing time per document at each review stage: human baseline [45, 46] versus VarEx pipeline.

| Stage | Human Baseline | Pipeline Runtime | Time Saved |
| --- | --- | --- | --- |
| Abstract Screening | 0.90 min/ref | 2 min 57 s (2.95 min) | −2.05 min/ref |
| Full-Text Screening | 7.00 min/ref | 3 min 12 s (3.20 min) | +3.80 min/ref |
| Data Extraction | 53.00 min/ref | 2 min 32 s (2.53 min) | +50.47 min/ref |
| **Total** | **60.90 min/ref** | **8.68 min/ref** | **+52.22 min/ref (85.7%)** |

In addition to time savings, LitScreen reduces downstream workload by excluding a large share of references at the abstract stage, saving computational cost and time for full-text review. On average, 60.8% of references were excluded during abstract screening, and this proportion varied by dataset. LitScreen excluded 94.1% of references for Education*→*Dementia, 86.9% for HTN*→*ADRD, and 48.1% for Bos et al. (2018).

## 4 DISCUSSION

This study demonstrates the potential of leveraging LLMs to automate and enhance the screening and data extraction process in systematic reviews and meta-analyses. By substantially reducing the time burden of citation screening while reliably retaining eligible studies, LitScreen helps ensure that systematic reviews are built on a more complete and timely evidence base. By addressing inconsistencies in confounder selection and patterns of omitted variable bias, VarEx can yield more reliable and comparable findings across studies.

### Contributions to Observational Studies

Our study makes two complementary contributions, LitScreen and VarEx. LitScreen is an automated screening pipeline, and VarEx is an automated extraction pipeline that, together, serve as a comprehensive resource for researchers conducting new observational studies or compiling evidence for systematic reviews. LitScreen substantially reduces the time burden of citation screening while reliably retaining eligible studies. By compiling data on reported confounders and identifying patterns of omitted-variable bias, VarEx aims to assist researchers in addressing inconsistencies in variable selection and their impact on effect estimates. VarEx provides researchers with a valuable reference point, enabling them to make informed decisions when selecting appropriate confounders for their studies. This, in turn, is expected to enhance the quality and reliability of epidemiological research. By addressing inconsistent confounder selection, VarEx advances epidemiological research methods and promotes the production of more reliable and comparable findings across studies.

## 5 Limitations and Future Work

LitScreen and VarEx hold great potential for advancing study screening and variable extraction in observational research. However, it is essential to address current limitations and implement strategic enhancements to fully realize this potential.

One limitation of our project, affecting both pipelines, was the restricted scope of our analysis, which limited our data to open-access papers in PubMed Central and Unpaywall. This constraint limited the breadth and depth of our dataset. Relying solely on published studies to identify relevant papers and extract variables may overlook highly relevant literature and less frequently reported variables. Moreover, even for papers whose full text is available, the quality of the XML parsing determines the coverage of the sections we can use. Some PMC XML files from older journals, or converted from pre-prints, often include malformed JATS markup, missing section labels, or combined Methods–Results blocks, which can lead the parser to misclassify or ignore sections that could be informative.

A further limitation affects both pipelines, but especially LitScreen, because the prompts must be manually modified for each new systematic review. No single prompt template works for every review, since each researcher may apply their own eligibility criteria, which keeps a human in the loop. Each of the study criteria, such as study design, exposure, and outcome, can differ in surface-level vocabulary across clinical domains through synonyms, abbreviations, and domain-specific terminology. The human expert must anticipate and explicitly handle each prompt set according to their criteria and the instructions provided in the tool. In addition, the adaptive semantic chunker has a fixed word-length ceiling of 500 words, which can fragment long methodological descriptions and degrade the passages available to both screening and extraction.

Finally, both screening and variable extraction are dependent on text generated during the retrieval stage. If the top-5 re-ranked chunks do not contain the passage most relevant to the criterion because of imprecise RAG queries, embedding distributional mismatch, or re-ranker scoring errors, then the full-text decision provides a wrong decision.

We also found inconsistencies in the outputs produced and the decisions made by large language models. The inconsistencies include, but are not limited to, making screening decisions based on incorrect evidence sentences, extracting variables that are not present in the input papers, and failing to adhere to the instructions provided. However, hallucination still remains an inevitable of LLMs[47]. Thus, this pipeline should be used as an assistant to a human expert rather than as a replacement for expert judgment.

A central direction for future work is the development of **SYNTHESISdb**, a standardized confounder database built on top of the variables extracted by VarEx. Future iterations should incorporate expert consultations and consensus-building exercises to address these gaps. This approach will help identify and incorporate critical yet underrepresented variables, thereby ensuring a more comprehensive database. Implementing causal feature selection methods will further expand the range of confounders, enhancing the database’s methodological rigor.

Expanding VarEx to include a broader array of risk factors, data sources, and research methodologies—such as meta-analyses, clinical trials, and longitudinal cohort studies—will provide a more comprehensive view of confounding variables and improve the generalizability of findings. Optimizing the user interface with intuitive search features, advanced data visualization tools, and clear guidelines will facilitate broader adoption and utility within the ADRD research community.

## 6 Conclusion

In conclusion, LitScreen and VarEx represent a significant step toward automating the screening and variable extraction tasks that underpin observational research. These pipelines can be used by researchers as assistants to identify eligible studies and to extract exposures, outcomes, and covariates from full-text studies. By automating screening and extraction and addressing inconsistencies in how study variables are identified and reported, LitScreen and VarEx support more reliable and comparable findings across studies. As an open-source and transparent solution, LitScreen considerably reduces screening time and effort while maintaining high accuracy, supporting rapid, reproducible, and scalable evidence synthesis for systematic review teams. VarEx’s structured output is designed to be compatible with existing tools and resources, such as ‘Metaconfoundr’, further enhancing its value and potential impact. As this research evolves, LitScreen and VarEx serve as a foundation for future work to develop SYNTHESISdb, a standardized confounder database, incorporate diverse data sources, and develop user-friendly interfaces and tools. Through these ongoing efforts, we strive to support the research community in conducting rigorous, reliable observational studies that can ultimately inform clinical practice and improve outcomes.

## Data Availability

All data produced are available online at: https://github.com/unmtransinfo/Screening
and
https://github.com/unmtransinfo/SYNTHESISdbGPT

https://github.com/unmtransinfo/SYNTHESISdbGPT

## Abbreviations

ADRD: Alzheimer’s Disease-Related Dementias
LLMs: Large Language Models
DAG: directed acyclic graph
HTN: Hypertension
LitScreen: Literature Screening Pipeline
VarEx: Variable Extraction Pipeline
SYNTHESISdb: Field specific Database for compiling information about confounding control reported in observational studies

## Acknowledgments

Funding from the US National Institutes of Health National Library of Medicine grant R00 LM013367 and National Institute of Mental Health grant R01MH129764 supported this research. The views expressed in this paper do not necessarily reflect the views of the National Library of Medicine of the National Institutes of Health.

## CRediT author statement

**Manjil Pradhan**: Methodology, Software, Formal Analysis, Writing - Original draft, Review & Editing, Data Curation, Visualizations **Rajesh Upadhayaya**: Software **Sarah Wenyon**: Conceptualization **Alexandria Viszolay**: Validation **Melissa Rethlefsen**: Conceptualization **Vincent Metzger**: Validation, Writing - Review & Editing **Gerardo Villarreal**: Validation **Santiago Alvarez Lesmes**: Validation **David Andrade**: Validation **Jason Timm**: Software **Scott A. Malec**: Conceptualization, Methodology, Software, Writing - Original draft, Review & Editing, Supervision, Project administrator, Funding acquisition

### Things To Do

#### Incomplete / placeholder content

- **Add LITSCREEN sub-study**: BPV screening study from COVIDENCE - describe at appropriate levels in intro, methods, results
- **Add LITSCREEN sub-study**: treatment for PTSD and self/harm study from COVIDENCE and describe at appropriate levels in intro, methods, results
- **Add VAREX substudy**: PTSD and self-harm/suicidality and describe at appropriate levels in intro, methods, results
- **Appendices:** Appendix A (MEDLINE query) and Appendix D (Screening Results) are empty headings.

**CRediT author statement Need to consult with Dr. Scott if we need this as it is based on journal**

## Appendix A: MEDLINE via OVID Search Query

This MEDLINE via OVID search strategy was developed with a health sciences librarian and run on 5 April 2024 against Ovid MEDLINE(R) ALL (1946 to 4 April 2024), returning 1,318 records. The strategy was derived from three prior published searches, the NICE dementia guideline NG97 (population terms), the SIGN observational studies MEDLINE filter, and the NICE hypertension guideline NG136. Table 7 lists the full strategy.

**Table 7:**
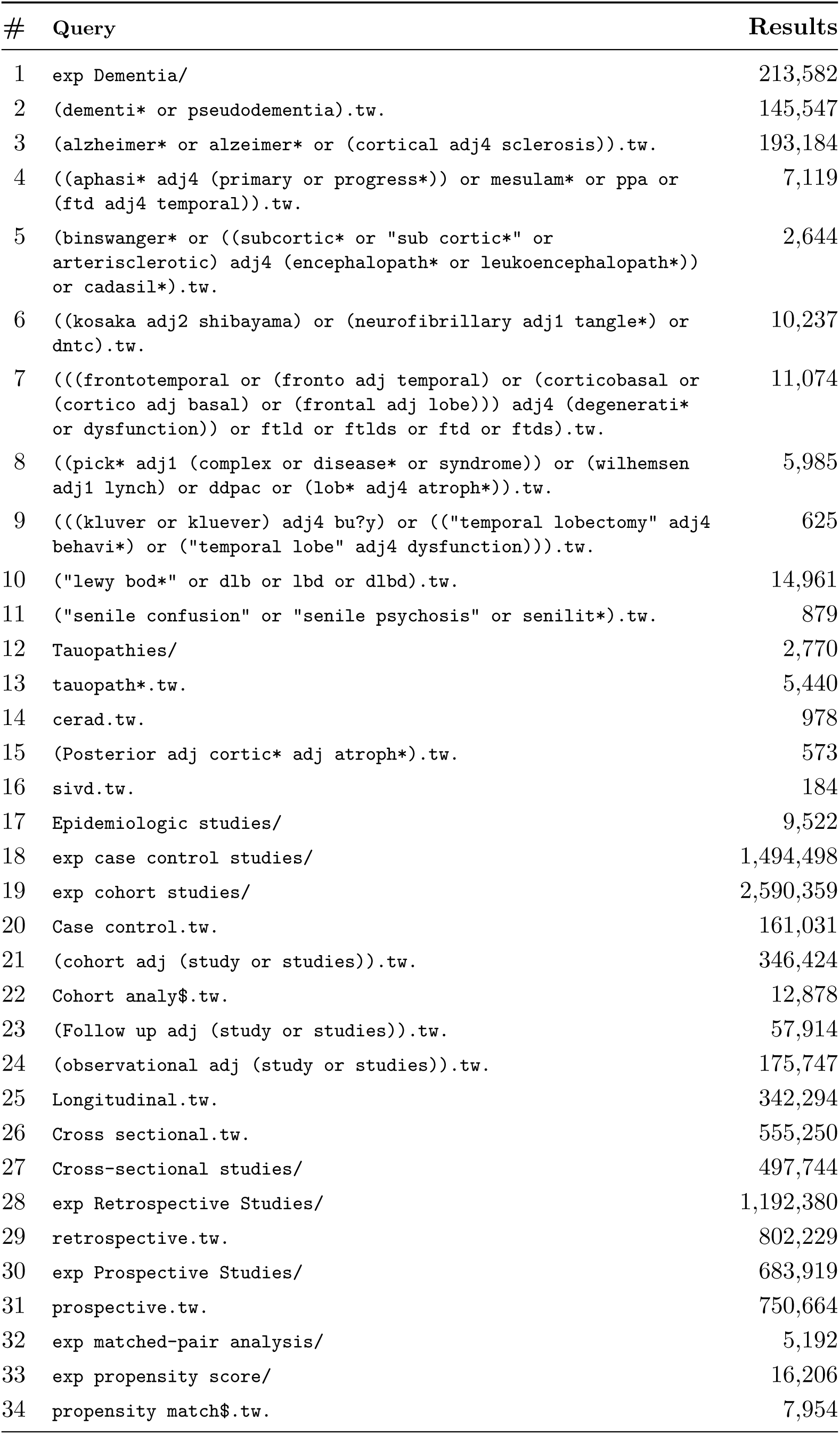
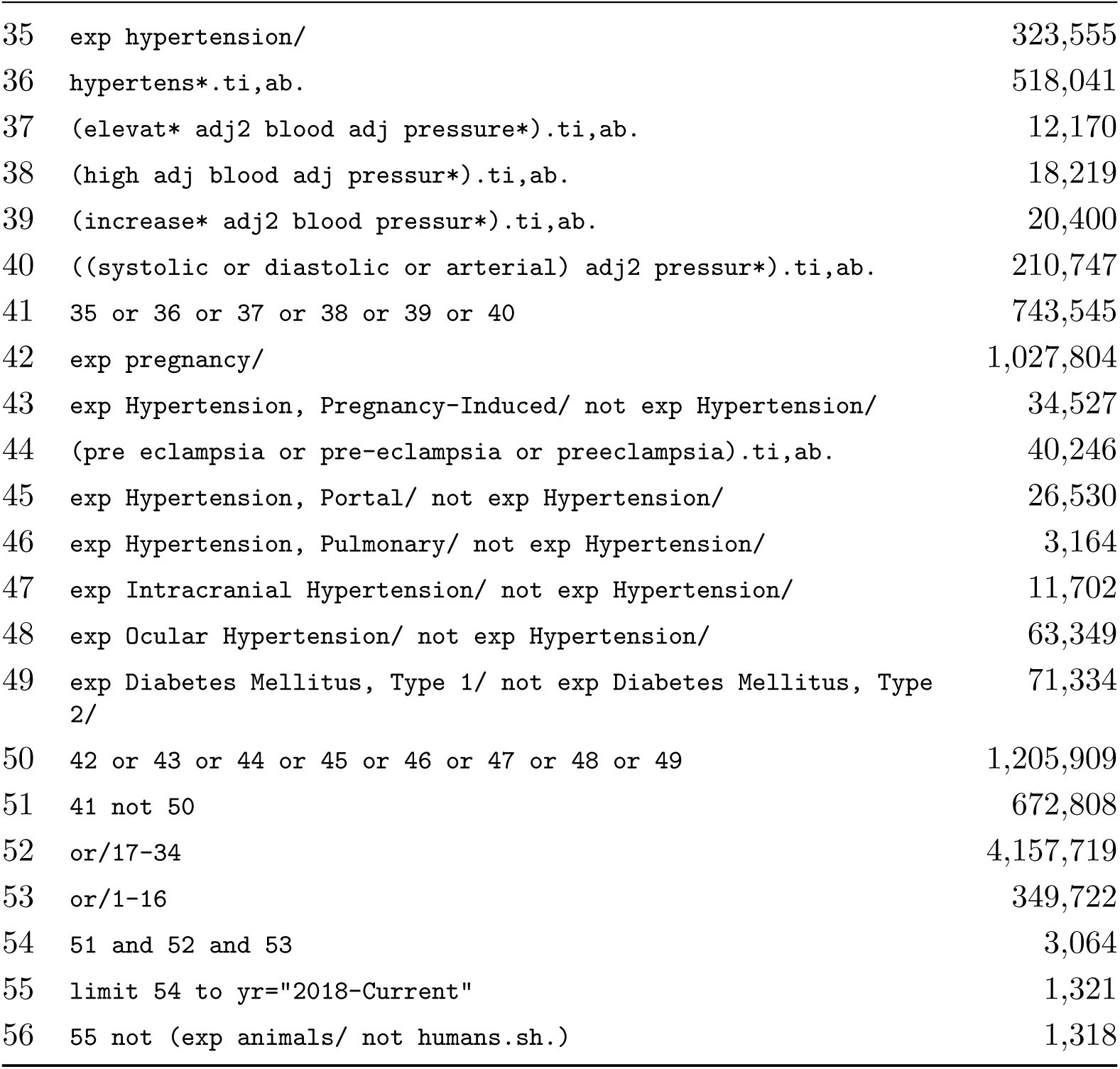
Ovid MEDLINE search strategy for dementia and hypertension.

## Appendix B: Abstract Evidence Extraction Prompt (Generic Template)

Phase 1 uses the same domain-neutral extraction prompt for all three criteria (Study Design, Exposure, Outcome); only the target term, description, definition, and few-shot examples change.

> You are a personal assistant to a biomedical researcher.

> Your task: identify and copy sentence(s) from the abstract that describe the study’s

> **[TARGET, e.g., EXPOSURE]** – [description of what this target means].

> **DEFINITION:** [criterion-specific definition of the target]

> Examples: [few-shot example sentences illustrating the target]

> ABSTRACT: [abstract text]

> Follow these steps carefully:

> **STEP 1 – IDENTIFY TOP N CANDIDATES**

> Select the top N sentences most likely to describe the **[TARGET]**. If fewer than N exist, list only those. Focus on sentences that directly name the [TARGET]. A sentence that lists multiple items or reports a risk estimate for a specific condition still counts – do NOT skip it because it also mentions other things.

> **STEP 1.5 – SELF-CHECK** (optional, criterion-specific) Ask yourself: [self-check question]. If YES for at least one candidate, continue to Step 2. If NO for all candidates, scan the abstract once more, looking specifically for sentences in results sections, sentences listing multiple conditions, and sentences reporting risk estimates or hazard ratios. Add any overlooked sentences to your candidate list before continuing.

> **STEP 2 – RELEVANCE CHECK**

> For each candidate ask: “Does this sentence directly describe the **[TARGET]**, or is it about something else (background, results, topic being studied)?” Write KEEP or REMOVE with a one-line reason for each candidate.

> **STEP 3 – VERBATIM CHECK**

> For each KEEP candidate, locate it in the abstract above and verify it appears word-for-word. Even one changed or missing word fails this check. Write VERIFIED or NOT VERBATIM for each.

> **FINAL_SENTENCES:**

> List only sentences that passed both Step 2 (KEEP) and Step 3 (VERIFIED), one per line, copied exactly from the abstract. If none survive, write: NOT STATED

### Appendix C: From Eligibility Criteria to Screening Prompts

This appendix traces how a published set of eligibility criteria becomes an executable screening prompt. We first reproduce the official criteria used as input (Section C.1), then show the three prompt-engineering strategies that operationalize them (Section C.2), and finally give the generic prompt templates those strategies produce (Section C.3). Each strategy is illustrated with a verbatim excerpt from the deployed prompts.

#### C.1 Official Eligibility Criteria (Input)

> “… the following inclusion criteria: (1) cohort studies or longitudinal studies conducted with routinely collected health care data (e.g., national medical registries or insurance databases); (2) determinant CHD (i.e., myocardial infarction with or without angina or coronary revascularization) or CHF; and (3) report of incident dementia diagnosis as the outcome (i.e., at least all-cause dementia or AD as its most common subtype).” [29]

#### C.2 Three Operationalization Strategies

A criterion stated in prose does not by itself tell a model how to read a confusable paper. We bridge that gap with three complementary strategies, each motivated by a recurring failure mode observed during development. Strategy (a) structures the reasoning, strategy (b) anchors it with worked examples, and strategy (c) hard-codes the boundary decisions that the model otherwise gets wrong.

##### (a) Chain-of-thought staged reasoning with self-reflection

Following chain-of-thought task decomposition [39] and self-refinement [48], every decision prompt forces a three-stage pass rather than an immediate verdict. The model first commits to an initial answer, then critiques it against criterion-specific reflection questions, then finalizes. This reduces impulsive exclusions that arise when a model answers before considering borderline cases. A representative reflection question from the hypertension exposure criterion:

> “Is hypertension mentioned only in an ‘adjusted for… ’ list (e.g., ‘adjusted for age, sex, hypertension, diabetes’) or only in the demographic/Table 1 description, with no analytic role? If yes, answer NO.”

##### (b) Few-shot contrastive examples

Reflection questions are paired with in-context worked examples [49], weighted toward contrastive counter-examples that mark exactly where a YES turns into a NO. These pin down the boundary far more reliably than an abstract definition. The deployed hypertension exposure prompt embeds this counter-example verbatim:

> “COUNTER-EXAMPLE (answer = NO): ‘Obesity was associated with dementia (HR 1.4). Subgroup analysis: among hypertensive participants, obesity was associated with dementia (HR 1.6).’ Here PRIMARY EXPOSURE = obesity, not hypertension — the hypertensive-subgroup HR is still measuring the obesity effect within a subgroup.”

##### (c) Rule-based encoding of recall traps

We inspected the model’s reasoning on boundary cases and, for each recurring error, added an explicit decision rule rather than leaving the judgment open, in line with evidence that itemized instructions make LLM decisions more accurate and consistent [50]. Most rules are *recall traps*: phrasings the model tended to read as disqualifying that should not exclude a paper. Operationally each criterion carries a *closed disqualifier list* alongside an explicit *not-a-disqualifier* list, as in the study-design criterion:

> “NOT disqualifiers — these REMAIN eligible (protect recall, answer YES): A **retrospective** registry/EHR cohort — it follows participants forward through time; it is longitudinal, NOT cross-sectional. **Record linkage** and **ICD-code identification** are cohort-building methods, NOT a systematic-review literature search.”

The same scaffold is reused across all nine validation datasets, each adding its own rules as errors surfaced. Table 8 lists representative error-to-rule pairs; the examples are illustrative rather than exhaustive. Shared across datasets are three meta-rules: an evidence-basis gate (DISQUALIFYING / CONFIRMING / INSUFFICIENT, where INSUFFICIENT can never exclude), a “not confirmed is not the same as violated” guard, and a requirement to quote the deciding sentence verbatim before judging.

**Table 8:** Representative error-to-rule pairs encoded as recall traps.

| Error the model made | Rule added | Dataset |
| --- | --- | --- |
| Counted BMI as an eligible outcome when it was only a covariate / adjustment variable | Role test: the measure must be a *target* related to hair GC, not analysis machinery | van der Valk |
| Excluded a cohort with a cross-sectional baseline arm as “longitudinal” | “A cross-sectional *analysis* is not a cross-sectional *study design*; a both-arms study is eligible via its cross-sectional arm” | van der Valk, Bos, Brouwer |
| Read a retrospective registry/EHR cohort as a literature review or as cross-sectional | “A retrospective registry/EHR cohort is longitudinal, not a systematic review → eligible” | Wolters, HTN |
| Tracked an already-diagnosed population as an incident-outcome cohort | “Participants already diagnosed at enrolment → NO” (genuine disqualifier) | Wolters, HTN, Education |
| Excluded dementia when it was one of several endpoints | “‘Listed alongside other endpoints’ is not a disqualifier” | Bos |
| Read population sampling as absence of trauma | “A community cohort that experienced a defined traumatic event is trauma-exposed; sampling is not the same as ‘no trauma’ ” | van de Schoot |
| Upgraded weak wording into a disqualifier | “‘Initially non-depressed’ is the remitted state, not first-onset → not a disqualifier” | Brouwer |

#### C.3 Resulting Prompt Templates (Output)

Combining the three strategies yields the generic templates below. Phase 2 (abstract) and Phase 3 (full text) share one structure; only the task description, criterion definition, context rules, reflection questions, and few-shot examples change per criterion.

##### Abstract decision prompt

> TASK: [criterion-specific task, e.g., “Determine whether a research paper meets the EXPOSURE criterion.”]

> **CRITERION:** *[criterion-specific definition]*

> *[criterion-specific context: qualifying and disqualifying rules]*

> **EXTRACTED SENTENCES:** *[verbatim sentences extracted in Phase 1]*

> **STAGE 1 – INITIAL ANALYSIS** *State an initial answer (YES / NO / UNCLEAR)*.

> **STAGE 2 – SELF-REFLECTION** *Critique the initial analysis against criterion-specific reflection questions*.

> **STAGE 3 – FINAL ANSWER** *Revise if reflection changed the view; otherwise confirm*.

> **HIGH-RECALL RULE.** *Absence of detail is NOT evidence of exclusion. Use NO only when the extracted sentences affirmatively state a disqualifying fact; otherwise answer UNCLEAR so the paper proceeds to full text*.

> **EVIDENCE-BASIS GATE.** *Declare DISQUALIFYING, CONFIRMING, or INSUFFICIENT; INSUFFICIENT forbids a NO*.

> *OUTPUT: INITIAL ANSWER, REFLECTION, [OUTPUT FIELD], REASONING, EVIDENCE BASIS, DISQUALIFIER VERBATIM, ANSWER, CONFIDENCE*.

##### Full-text decision prompt

> TASK: [criterion-specific task] **CRITERION:** [criterion-specific definition] [criterion-specific context]

> **FULL-TEXT EVIDENCE (Methods / Results):** [retrieved chunks]

> **STAGE 1 – INITIAL ANALYSIS** Commit to YES or NO; UNCLEAR is not permitted when full-text evidence is available.

> **STAGE 2 – SELF-REFLECTION** Critique against criterion-specific reflection questions.

> **STAGE 3 – FINAL ANSWER** Answer YES or NO only.

> OUTPUT: INITIAL ANSWER, REFLECTION, [OUTPUT FIELD], REASONING, ANSWER, CONFIDENCE.

### Appendix D: LitScreen Phase 3 Criterion-Specific RAG Retrieval Queries for HTN *→* ADRD

For Phase 3 full-text verification, each unresolved criterion is matched against the Methods and Results sections of the paper using a criterion-specific retrieval query. The query is embedded with bioLinkBERT-large and used to retrieve the top-50 candidate chunks via cosine similarity and Maximal Marginal Relevance (MMR), which are then reranked to the top-5 chunks using a cross-encoder (BAAI/bge-reranker-large).

#### Study Design Query

> This was a prospective population-based cohort study with longitudinal follow-up. Participants were community-dwelling adults without dementia at baseline enrolled from registries. Retrospective cohort study using electronic health records linked to administrative databases. Eligible participants were followed over time for incident dementia outcomes. This epidemiologic study enrolled participants and followed them prospectively for cognitive outcomes.

#### Exposure Query

> Hypertension was the primary exposure of interest in this study. Systolic blood pressure (SBP) and diastolic blood pressure (DBP) were measured at baseline and followup. We examined whether high blood pressure predicts incident dementia or cognitive decline. Antihypertensive treatment use and risk of Alzheimer disease and dementia. Blood pressure was classified as hypertensive if SBP ≥140 mmHg or DBP ≥90 mmHg. Midlife hypertension and late-life blood pressure as predictors of dementia risk.

#### Outcome Query

> Incident dementia was the primary outcome ascertained during follow-up. New cases of dementia and Alzheimer disease were identified using DSM or ICD criteria. Mild cognitive impairment (MCI) and Alzheimer’s Disease and Related Dementias (ADRD) were assessed. Hazard ratio for incident all-cause dementia was estimated using Cox proportional hazards regression. Participants were followed until dementia diagnosis, death, or end of study.

### Appendix E: VarEx Role-Specific RAG Retrieval Queries

For each role, the query below is embedded with NeuML/pubmedbert-base-embeddings and used to retrieve candidate chunks via embedding similarity, which are then re-ranked with the BAAI/bge-reranker-large cross-encoder to select the top four chunks per role supplied to the extraction LLM. Unlike LitScreen’s criterion-specific queries, these templates are domain-general and designed to match how exposures, outcomes, and covariates are reported across biomedical papers generally, rather than to any single exposure-outcome pair.

#### Exposure Query

> Participants were classified according to. was measured using a self-reported questionnaire at baseline. Participants were categorized into groups based on. was stratified as follows. The independent variable was calculated as.

#### Outcome Query

> Cases were ascertained through clinical evaluation. was assessed using standardized neuropsychological tests. was diagnosed according to established criteria. The dependent variable was defined as. was identified through medical records and clinical assessment.

#### Covariate Query

> The regression model adjusted for the following variables. Variables included in the multivariable model were. The analysis was adjusted for. Models were additionally adjusted for. The model controlled for. These variables were covariates in all models. Levels were corrected for age and gender. The independent variables included in the model were. Variables were entered simultaneously into the regression model.

### Appendix F: Screening Results

#### Screening Performance on SYNERGY Benchmark

Table 9 reports screening classification performance on the labeled SYNERGY benchmark datasets. Because the production corpora (Hypertension*→*ADRD and Education*→*Dementia) lack ground-truth inclusion labels, screening performance on those datasets was evaluated manually by domain experts and is not reported here.

**Table 9:**
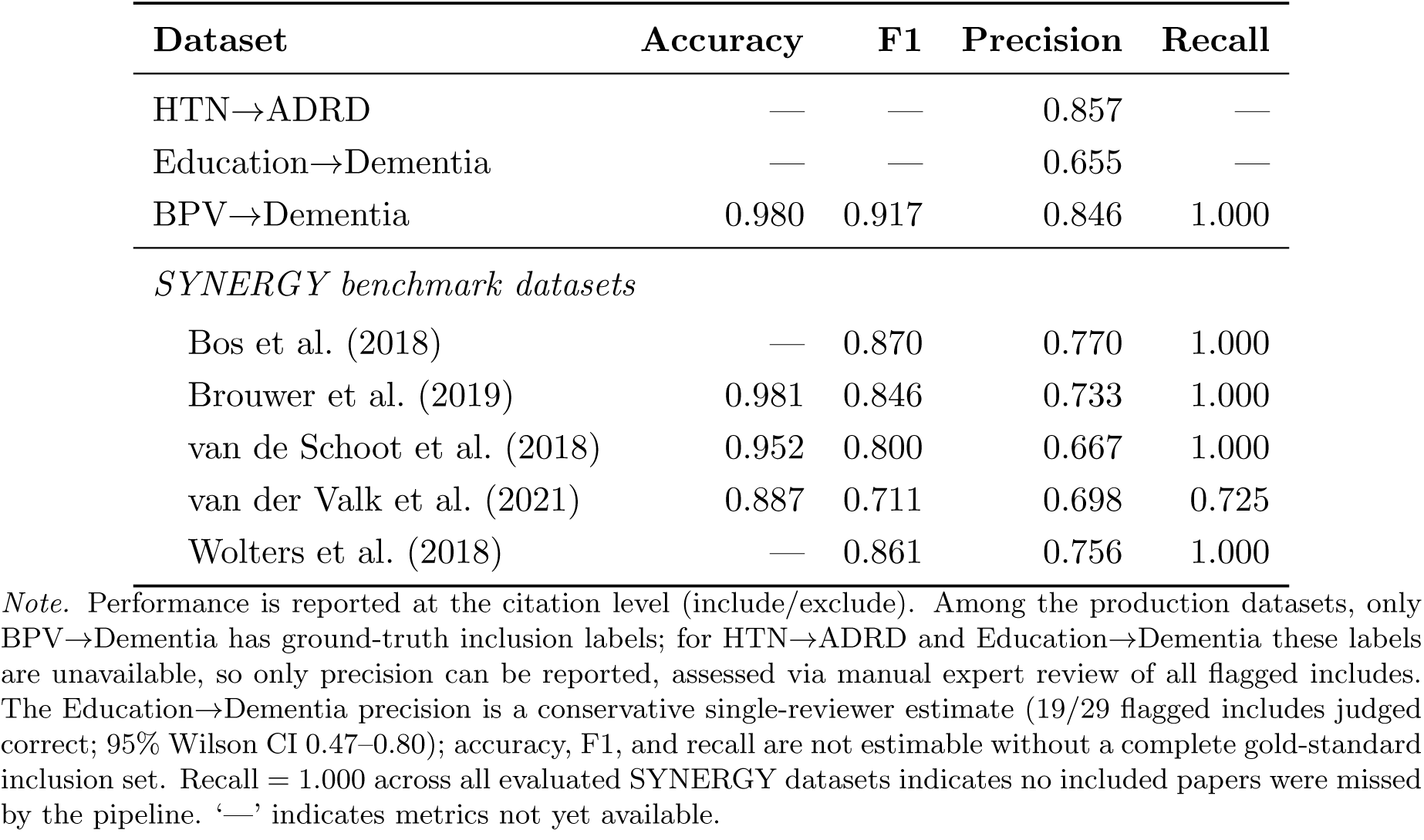
Screening performance across production and SYNERGY benchmark datasets.

| Dataset | Accuracy | F1 | Precision | Recall |
| --- | --- | --- | --- | --- |
| HTN→ADRD | — | — | 0.857 | — |
| Education→Dementia | — | — | 0.655 | — |
| BPV→Dementia | 0.980 | 0.917 | 0.846 | 1.000 |
| *SYNERGY benchmark datasets* |  |  |  |  |
| Bos et al. (2018) | — | 0.870 | 0.770 | 1.000 |
| Brouwer et al. (2019) | 0.981 | 0.846 | 0.733 | 1.000 |
| van de Schoot et al. (2018) | 0.952 | 0.800 | 0.667 | 1.000 |
| van der Valk et al. (2021) | 0.887 | 0.711 | 0.698 | 0.725 |
| Wolters et al. (2018) | — | 0.861 | 0.756 | 1.000 |

*Note.* Performance is reported at the citation level (include/exclude). Among the production datasets, only BPV→Dementia has ground-truth inclusion labels; for HTN→ADRD and Education→Dementia these labels are unavailable, so only precision can be reported, assessed via manual expert review of all flagged includes. The Education→Dementia precision is a conservative single-reviewer estimate (19/29 flagged includes judged correct; 95% Wilson CI 0.47–0.80); accuracy, F1, and recall are not estimable without a complete gold-standard inclusion set. Recall = 1.000 across all evaluated SYNERGY datasets indicates no included papers were missed by the pipeline. ‘—’ indicates metrics not yet available.

### Appendix H: Covariate-Matching Threshold Sensitivity Analysis

A matched pair was accepted as a true positive only if its cosine similarity met or exceeded the chosen threshold; pairs below the threshold were scored as a false negative (missed reference covariate) and a false positive (spurious extracted covariate). We swept this threshold from 1.0 to 0.4 across the full 10-paper reference standard (101 reference covariates) and recomputed aggregate precision and recall at each value (Table 10, Figure 5).

**Figure 5:**
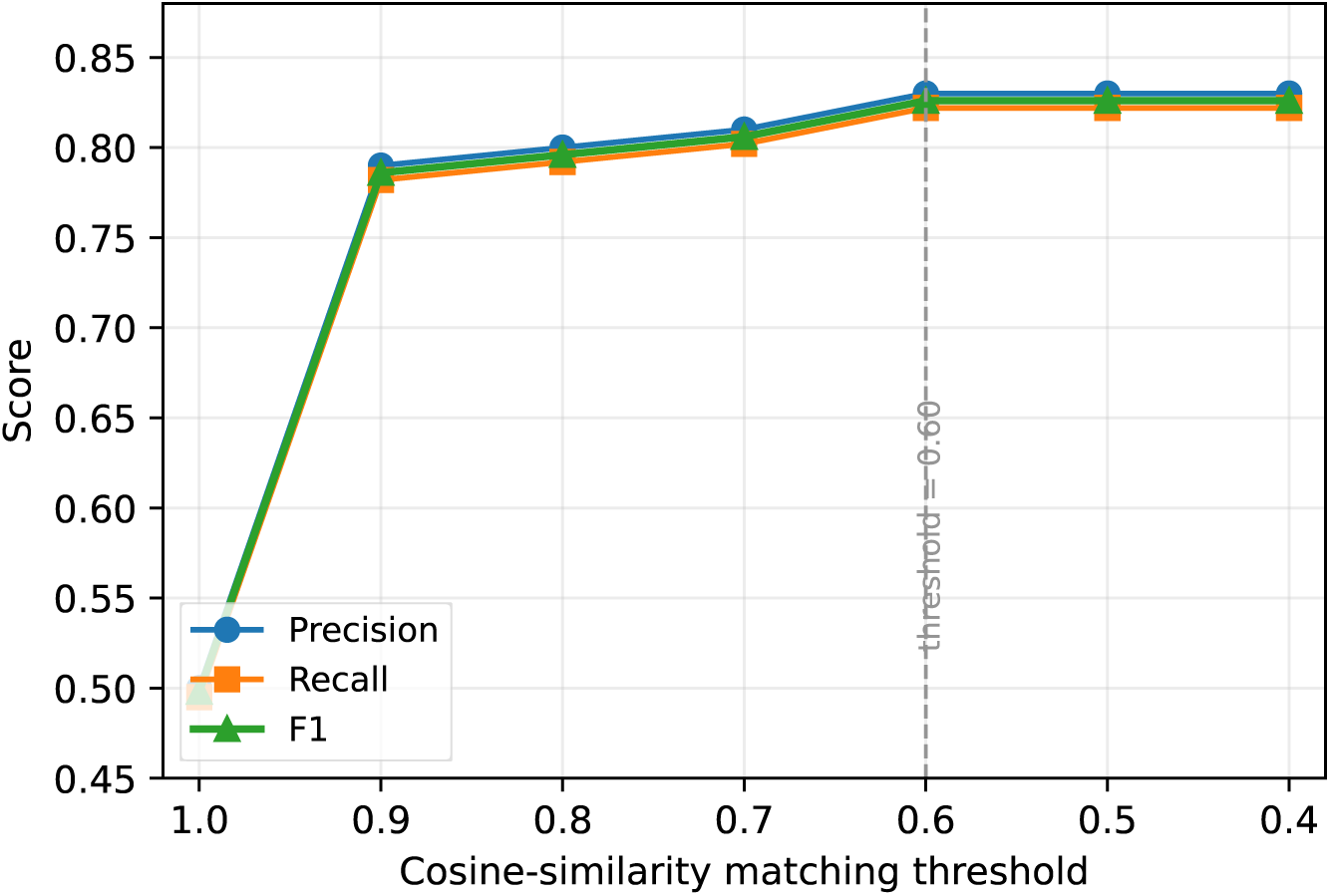
Precision, recall, and F1 as a function of the covariate matching threshold, computed on the full 10-paper reference standard. Performance plateaus for thresholds at or below 0.60.

**Table 10:** Aggregate matching performance across the 10-paper reference standard at decreasing similarity thresholds.

| Threshold | TP | FP | FN | Precision | Recall |
| --- | --- | --- | --- | --- | --- |
| 1.00 | 50 | 50 | 51 | 0.500 | 0.495 |
| 0.90 | 79 | 21 | 22 | 0.790 | 0.782 |
| 0.80 | 80 | 20 | 21 | 0.800 | 0.792 |
| 0.70 | 81 | 19 | 20 | 0.810 | 0.802 |
| **0.60** | **83** | **17** | **18** | **0.830** | **0.822** |
| 0.50 | 83 | 17 | 18 | 0.830 | 0.822 |
| 0.40 | 83 | 17 | 18 | 0.830 | 0.822 |

Requiring near-exact similarity (threshold *≥* 0.9) penalizes covariates that are semantically identical but phrased differently (e.g., “BMI” versus “body mass index”), driving precision and recall down to roughly chance levels. Performance rises sharply as the threshold is relaxed from 1.0 to 0.6, then plateaus: aggregate performance is identical at every threshold from 0.65 down to 0.40, because no covariate pair in this corpus has a similarity score in that band. We therefore did not select 0.60 because it uniquely maximizes performance on this corpus; rather, it is the most conservative (strictest) threshold within the empirically optimal plateau. Choosing the upper edge of the plateau, rather than its more permissive lower edge, reduces the risk that the threshold is overfit to this 10-paper corpus and would admit spurious matches on covariates outside it.

To illustrate why the plateau exists, we constructed a small illustrative example of 9 reference/extracted covariate pairs describing the same construct with different wording (e.g., “Sex” vs. “Gender,” “Diabetes mellitus” vs. “Diabetic status”), plus one extracted term with no true counterpart in the reference list (“Marital status”), run through the same matching procedure. This example is illustrative, not a statistical sample: at a threshold of 1.0, none of the 9 paraphrase pairs match, since synonymous covariate names rarely share identical embeddings; at 0.60, 8 of the 9 pairs match correctly, and the unrelated term is correctly never matched at any threshold down to 0.40. The one pair missed at 0.60 (“Smoking status” vs. “Tobacco use,” similarity 0.584) is recovered at 0.50, consistent with the corpus-level plateau observed above.

